# Linear predictive coding electroencephalography algorithms predict Parkinson’s disease mortality using out-of-sample tests

**DOI:** 10.1101/2025.07.07.25331047

**Authors:** Simin Jamshidi, Arturo I. Espinoza, Jonathan T. Heinzman, Patrick May, Ergun Y. Uc, Nandakumar S. Narayanan, Soura Dasgupta

**Affiliations:** Department of Computer and Electrical Engineering, College of Engineering, University of Iowa, Iowa City, IA; Department of Neurology, Carver College of Medicine, University of Iowa, Iowa City, IA; Broadlawns Medical Center, 1801 Hickman Rd, Des Moines, IA 50314; NVIDIA, Westford, MA; Neurology Service, Veterans Affairs Medical Center, Iowa City, IA

**Keywords:** Parkinson’s Disease, Mortality Prediction, Machine Learning, Linear Predictive Coding

## Abstract

**Background:** Parkinson’s disease (PD) increases mortality is difficult to predict because of its heterogeneity and the availability of very few reliable which prognostic markers.

**Objectives:** We used electroencephalography (EEG) and the Linear Predictive Coding EEG Algorithm for PD (LEAPD) for binary classification of 3-year mortality status and correlation between LEAPD indices and time to death.

**Methods:** 2-minutes resting-state EEG from 94 PD patients (59 channels, 22 deceased within 3 years of recording) was used for binary classification of 3-year mortality status. Single-channel classification using a balanced dataset of 44 was performed using leave-one-out cross-validation (LOOCV). Robustness was evaluated by truncating the recordings. LOOCV Spearman’s correlation coefficient (ρ) was obtained between LEAPD indices and time to death. Optimum hyperparameters obtained from a balanced training dataset of 30 were tested on the remaining 64 patients by 10,000 randomized comparisons of 7 vs 7, using 5 channel combinations Hyperparameters for the best ρ, using the same training dataset were for the out-of-sample correlation for the remaining 7 deceased.

**Results:** In LOOCV analysis several channels yielded 100% accuracy with robust performance from five. The correlations ranged between ρ =-0.59 to-0.86; were significant after adjusting for age, cognitive and motor impairment. Out-of-sample testing using the best-performing 5-channel combination yielded a mean accuracy of 83%. Out-of-sample Spearman’s ρ was-0.82.

**Conclusion:** LEAPD provides a robust approach for binary classification of mortality in PD from resting-state EEG. LEAPD indices correlate with survival duration, independent of clinical predictors, suggesting potential utility as a continuous neurophysiological biomarker.

## Introduction

Parkinson’s disease (PD) is the second most common neurodegenerative disorder worldwide, with its prevalence increasing due to aging populations and modern environmental exposures (1,2). While motor symptoms—such as bradykinesia, tremor, rigidity, and postural instability—are hallmark features, non-motor symptoms like cognitive decline and autonomic dysfunction also contribute significantly to morbidity and mortality (3–5). PD patients face an elevated mortality risk, with clinical milestones such as hallucinations, falls, dementia, and nursing home placement markedly increasing the likelihood of death (6) A systematic review and meta-analysis estimated a pooled mortality ratio in PD of approximately 1.5, with survival declining by about 5% for each year of follow-up (7). Accurately determining mortality risk in PD can guide clinical decision-making, support advance care planning, and help identify patients who may benefit from targeted interventions.

Despite this need, objective biomarkers for predicting mortality in PD remain scarce (8–10). Clinical features such as motor impairment and gait dysfunction are associated with increased mortality risk (11), and non-motor symptoms—such as psychosis, cognitive impairment, and autonomic issues—have also been linked to worse survival outcomes (12). However, although clinical metrics of motor (e.g., Unified Parkinson’s Disease Rating Scale [UPDRS] III scores) and non-motor (e.g., Montreal Cognitive Assessment [MoCA] scores) function can offer some information about mortality risk and overall disease burden, they are not sufficiently accurate (11,13).

Our group has recently developed neurophysiological markers that predict clinical aspects of PD using scalp-recorded electroencephalography (EEG) (3,14). EEG-based biomarkers have shown promise for detecting both motor and cognitive dysfunction in PD, with resting-state EEG in particular being sensitive to disease-related cortical changes (3,8,15,16). For instance, spectral EEG features have been shown to correlate with MoCA scores and other cognitive measures (3,14). In contrast to these prior studies, our approach focuses specifically on mortality prediction using the Linear Predictive Coding EEG Algorithm for PD (LEAPD).

LEAPD was originally developed by Anjum et al. (2020) as a computationally efficient method that uses linear predictive coding to transform EEG time-series signals into spectral features, enabling classification of PD patients versus non-PD controls (9). LEAPD is designed to maximize accuracy while using the fewest number of EEG channels—a key advantage in clinical settings where fast setup, patient comfort, and feasibility are essential—making it well suited for real-time applications (9,17).

In this study, we tested the hypothesis that LEAPD can predict mortality in PD using resting-state EEG recordings (2 minutes, 64 channels; 59 retained for analysis after artifact rejection). We conducted two sets of experiments.

The first set of experiments aimed to determine whether EEG recordings could distinguish patients who would die within three years from those who would survive—a binary classification task—and consisted of two steps: (1) evaluating single-channel classification performance using cross-validation, and (2) testing the generalizability of the classifier in an independent out-of-sample dataset.

In the first step, we performed single-channel classification, where each of the 59 EEG channels was analyzed independently. This approach allowed us to identify which channels carried the most informative signal for distinguishing deceased from living patients. Using a balanced dataset of 44 PD patients (22 deceased within 3 years of EEG recording, 22 living), we applied leave-one-out cross-validation (LOOCV)—a method where the model is trained on all but one subject and tested on the held-out subject, repeated for every individual. This provided a robust estimate of classification accuracy across the group.

To test whether accurate classification could be maintained with less data, we conducted a truncation analysis in which EEG recordings were shortened by systematically removing data from the end of each 2-minute segment. This simulates real-world scenarios where shorter recordings may be more practical or necessary. The goal was to assess the robustness of the classifier when less data is available.

In the second step, we evaluated the generalizability of the LEAPD classifier using an out-of-sample test. A training dataset of 30 patients (15 deceased, 15 living) was used to optimize key hyperparameters—settings that control how the LEAPD algorithm processes data, including frequency band, Linear Predictive Coding (LPC) order, and hyperplane dimension. These hyperparameters were optimized using the full EEG recordings (e.g., not truncated) across all the 59 usable channels.

To assess performance beyond the training sample, we applied the optimized classifier to an independent test set of 64 patients (7 deceased, 57 living). To ensure balanced comparisons, we generated 10,000 random test sets of 7 deceased vs. 7 living patients. Both single-channel and multi-channel LEAPD classifiers were evaluated. In the multi-channel framework, a classifier integrates information from several EEG channels (e.g., 2 to 5 channels), with the final decision made based on the geometric mean of LEAPD indices from those channels. This approach allowed us to test whether combining data from multiple informative regions improves prediction compared to single-channel classification alone.

In the second set of experiments, we asked whether the LEAPD index could serve as a continuous biomarker—that is, not only distinguishing whether a patient died within three years, but also correlating with how soon death occurred. We calculated Spearman’s rank correlation coefficients ρ between LEAPD indices and time to death in the subset of 22 deceased patients. A strong negative correlation would suggest that higher LEAPD indices are associated with shorter survival, reflecting proximity to death.

To test generalizability, we optimized hyperparameters using a training subset of 30 patients (15 deceased, 15 living), then applied those parameters in an out-of-sample correlation analysis on the remaining 7 deceased patients, evaluating the association between their LEAPD indices and time to death.

Our findings show that LEAPD can accurately classify mortality status from short EEG recordings and that LEAPD indices are correlated with survival duration, supporting their use as both binary and continuous neurophysiological biomarkers of mortality risk in PD (3,9).

## Methods

### Participants

We recruited a total of 94 PD patients from the University of Iowa Movement Disorders Clinic between 2017 and 2021. All patients were examined by a movement disorders physician to verify that they met the diagnostic criteria recommended by the United Kingdom PD Society Brain Bank diagnostic criteria. All PD patients were determined to have the decisional capacity to provide informed consent in accordance with the Declaration of Helsinki and the Ethics Committee on Human Research. We obtained written informed consent from each participant. All research protocols were approved by the University of Iowa Human Subjects Review Board (IRB# 201707828). Data from some of these patients have been published previously (3,18).

All EEG recordings of PD patients were performed in the medication “ON” state to enhance clinical applicability. Initial training and validation datasets were analyzed prior to completing EEG recordings in 2021. The date of death for deceased participants was determined using comprehensive chart review from 2023–2024, and data analyses were completed in 2025.

### Study Design

The study consisted of two sets of experiments where the first set of experiments focused on using resting-state EEG to classify PD patients by three-year survival status, following a two-step binary classification process.

First, to evaluate whether EEG signals from individual scalp channels could predict three-year mortality in PD patients, we performed single-channel classification using a balanced cohort of 44 PD patients, consisting of 22 living and 22 deceased patients. Classification performance was assessed using LOOCV. To evaluate the robustness of the classifier and to assess performance under reduced data conditions, we conducted a truncation analysis, in which the length of EEG data available for each subject was systematically reduced. Specifically, classification accuracy was tested at four levels of data retention (100%, 90%, 66%, or 50%), to simulate varying degrees of data availability and to examine the stability of LEAPD under these constraints.

Second, to assess the accuracy in binary 3-year mortality classification of LEAPD on new, unseen data not used in training, we conducted an out-of-sample evaluation. A training dataset of 30 PD patients (15 deceased, 15 living) was used to optimize LEAPD hyperparameters. These optimized hyperparameters were applied to an independent test cohort of 64 PD patients (7 deceased, 57 living), ensuring a strict separation between the training and testing phases. To create balanced test sets, we fixed the 7 deceased patients and randomly sampled 7 living patients from the pool of 57, repeating this process 10,000 times to generate distinct 7-vs-7 comparisons. Classification performance was evaluated using a multi-channel LEAPD classifier, applied to combinations of 5 EEG channels. The mean and standard deviation of classification accuracy were computed across these 10,000 trials to quantify the model’s generalizability under realistic variability in the test data.

The second set of experiments aimed to evaluate whether LEAPD indices could serve as a continuous biomarker of survival duration in PD.

First, we computed LEAPD indices for each of the 22 deceased patients in the 44-subject dataset previously used for single-channel classification. For each subject, we used a leave-one-out cross-validation framework: in each iteration, one deceased subject was excluded, and the LEAPD hyperplanes were constructed using the remaining 21 deceased and all 22 living subjects. We then correlated the resulting LEAPD indices with the number of days between EEG acquisition and death. Spearman’s ρ was calculated separately for each of the 59 usable EEG channels, enabling identification of those channels most strongly associated with survival duration. The hyperparameter settings that yielded the most negative Spearman’s ρ values were selected for further.

Second, a balanced dataset of 15 deceased and 15 living patients was used to identify hyperparameter settings that produced the most negative Spearman’s ρ values, indicating stronger inverse associations between LEAPD indices and time to death. These values reflect that higher LEAPD indices are associated with shorter survival durations, supporting their potential as a continuous prognostic marker. This analysis built upon the same classification framework described earlier. Then, out-of-sample tests were conducted to compute Spearman’s correlation between LEAPD indices and time to death for the remaining 7 deceased patients from the full cohort of 94 PD patients.

### EEG Data Collection and Preprocessing

Resting-state EEG signals were recorded using a 64-channel actiCAP system, (Brain Products GmbH, Gilching, Germany), sampled at 500 Hz. The Pz electrode was used as a reference and excluded from analysis. Noisy or artifact-laden channels (TP9, TP10, FT9, and FT10) were identified and removed using the FASTER pipeline and the pop_rejchan function in EEGLAB (19).

### LEAPD and Feature Extraction to determine the LEAPD Index

LEAPD is an EEG-based signal processing method developed to distinguish patients with PD from healthy controls using resting-state EEG data. It encodes EEG time series into efficient feature representations, enabling fast, real-time classification (12). In this study, we adapted LEAPD to predict mortality in PD patients by identifying patterns in their EEG signals. This adaptation allowed LEAPD to differentiate between patients who survived more than three years after EEG acquisition and those who did not.

Filtered EEG data from each of the 94 PD patients were encoded into LPC vectors using linear predictive coding. An *n*-th order LPC vectors represents each sample of a time series as a weighted sum of the previous *n* samples. The coefficients a_l_ that minimize the Mean Square Error (MSE) are the LPC coefficients, as depicted in Figure 1c. The LPC coefficients encode the filtered EEG signal. These coefficients were then centered by subtracting their mean, resulting in LPC feature vectors.

**Figure 1.**
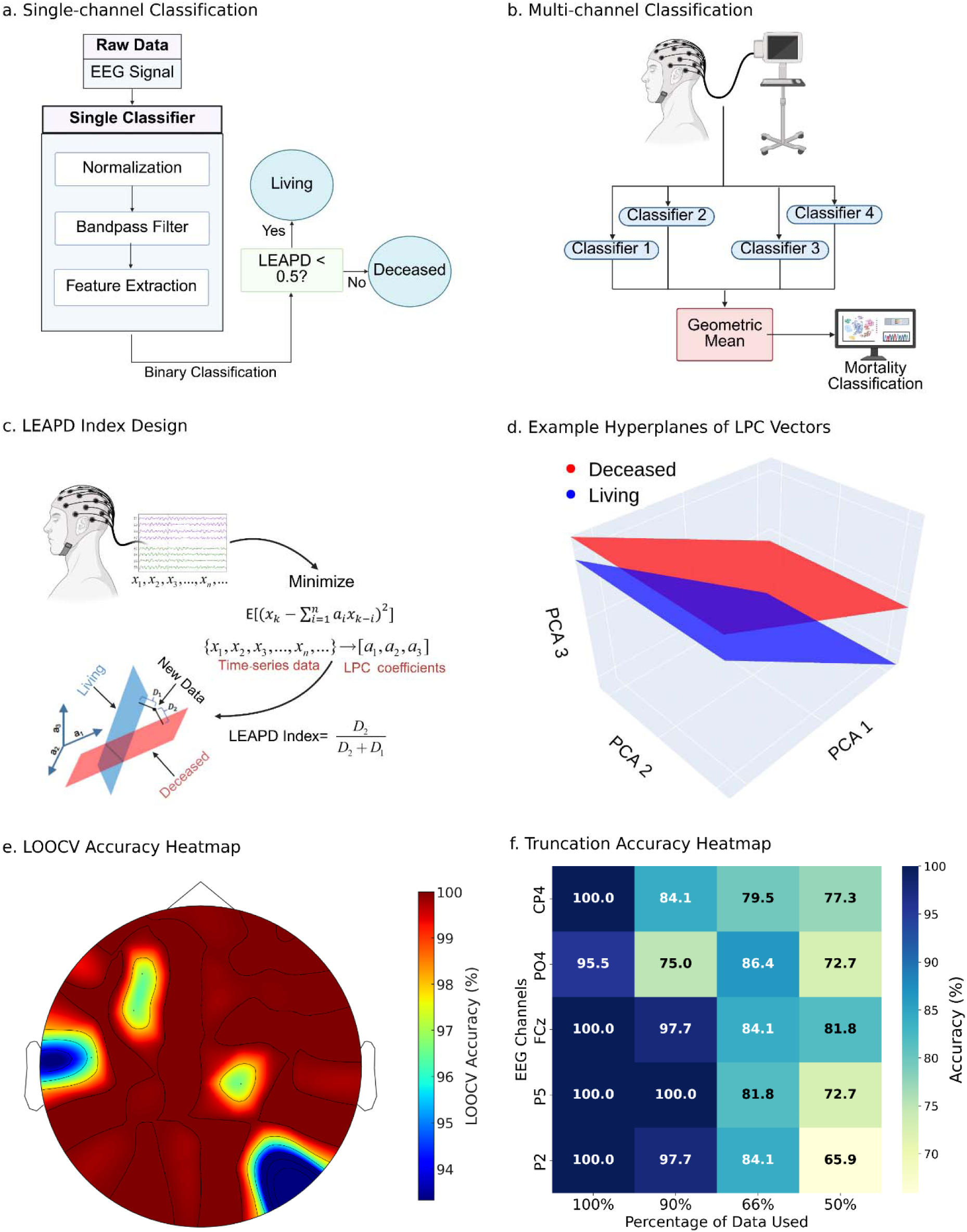
LEAPD workflow, LOOCV, and truncation analysis. **(a)** Single-channel classification pipeline with preprocessing and LEAPD-based binary decision. **(b)** Multi-channel classification using the geometric mean of all the classifiers trained on individual EEG channels. **(c)** LEAPD index design using LPC vectors to encode EEG and compute Euclidean distances to “Deceased” and “Living” hyperplanes. **(d)** Principal component analysis (PCA) visualization of example LPC coefficient hyperplanes showing the distance of separation between the Deceased and Living groups. **(e)** Topographic heatmap of LOOCV classification accuracy across EEG channels. **(f)** Accuracy heatmap for the top 5 EEG channels (CP4, PO4, FCz, P5, and P2) across 4 data truncation levels (100%, 90%, 66%, 50%).

In this study, we assumed that LPC vectors from the deceased and living patient groups lie on separate m-dimensional hyperplanes. These hyperplanes were identified using principal component analysis (PCA), which defines the geometric subspace spanned by each group’s LPC vectors. Figure 1c illustrates this conceptualization, and Figure 1d shows an example with m= 3. Using EEG recording data from each PD patient, we compute the distances D_l_ and D_2_ of the LPC vectors from the two hyperplanes representing data from living or deceased PD patients, respectively. The LEAPD index D_l_/(D_l_ + D_2_) is then computed for each PD patient. EEG data from PD patients that died within 3 years after the EEG recording were classified by a LEAPD index value of 0.5 or more.

A training phase is required to identify optimal hyperparameters and train the LEAPD model for both LOOCV and out-of-sample testing. In this study, the training dataset was used to tune the LEAPD model by selecting the best hyperparameter settings and deriving the representative hyperplanes for the living and deceased groups. The following hyperparameters were systematically optimized: bandpass filter range (0.1– 100 Hz), LPC order (2–10), the dimension of the hyperplanes, and optimal EEG channel selection. The best-performing. In this study, training data were used to identify optimized hyperparameters and the two hyperplanes. Hyperparameter combinations were identified by maximizing classification accuracy. For multi-channel analyses, a single LEAPD index was computed as the geometric mean of the indices from each individual channel.

### Single-Channel Classification and Truncation Analysis

We first evaluated the discriminative power of individual EEG channels to classify PD patients who died within three years (deceased) vs those who survived (living). A balanced dataset of 44 PD patients (22 deceased, 22 living) was used. LOOCV was applied to compute the LEAPD index for each subject and assess classification performance across all usable EEG channels (59 total, excluding Pz, TP9, TP10, FT9, and FT10). All channels yielded high classification accuracy, with many reaching 100%. These results are visualized as a scalp topographic heat map in Figure 1e.

To test classifier robustness under conditions of limited data availability, we performed a truncation analysis on the same 44-subject dataset. EEG data were systematically truncated to 90%, 66%, and 50% of their original duration, simulating scenarios with incomplete or noisy recordings. Truncation was applied from the end of the recordings. At each truncation level, classification performance was evaluated using the previously optimized hyperparameters without retraining. This analysis tested the resilience of LEAPD under realistic constraints such as shorter or interrupted EEG recordings in clinical settings.

### Multi-Channel Classification and Out-of-Sample Validation

To assess the generalizability of LEAPD to unseen data, we performed out-of-sample validation. A balanced training dataset of 30 PD patients (15 deceased, 15 living)— drawn from the original 44-subject set—was used to optimize hyperparameters. These settings were then applied to an independent test cohort of 64 PD patients (7 deceased, 57 living), ensuring no subject overlap between training and test sets.

To create balanced test comparisons, we fixed the 7 deceased patients and randomly sampled 7 living patients from the remaining pool. This process was repeated 10,000 times, generating distinct 7-vs-7 datasets. Multi-channel LEAPD classification (using the best-performing 5-channel combination) was applied to each test set. Classification accuracy, sensitivity, specificity, and AUC were averaged across the 10,000 out-of-sample trials, quantifying model performance and stability under realistic data variability.

### Correlation Analysis Between LEAPD Indices and Survival Duration

To evaluate the LEAPD index as a continuous biomarker for survival duration, we computed correlations between the index and time to death. Using the same 44-subject dataset (22 deceased, 22 living) from the single-channel classification analysis, we implemented an LOOCV framework. For each deceased subject, their data were excluded, and separate deceased and living hyperplanes were constructed using the remaining 21 deceased and all 22 living patients. The resulting LEAPD index (unthresholded, ranging from 0 to 1) for the excluded deceased subject was then correlated with their survival duration (i.e., days from EEG recording to death).

This procedure was repeated independently across 59 EEG channels, and Spearman’s rank correlation coefficient ρ was computed for each. Channels were ranked by the magnitude of negative correlations, where more negative *p* values indicated a stronger inverse relationship between LEAPD index and survival duration—suggesting that higher LEAPD indices reflect shorter life expectancy.

Statistical significance was assessed using p-values for Spearman’s *p*, and partial correlations were also computed to adjust for potential confounds such as age, MoCA scores, and UPDRS-III motor scores.

Finally, an out-of-sample correlation analysis was performed. A balanced training dataset of 30 PD patients (15 deceased, 15 living) was used to identify hyperparameters that yielded the most negative Spearman’s *p* values. These optimized settings were then applied to compute LEAPD indices for the remaining 7 deceased patients (from the full cohort of 94 PD patients). Spearman’s correlation was again calculated between the LEAPD indices and time to death to evaluate the model’s prognostic potential in unseen subjects.

For the classification and correlation analyses, separate LEAPD indices were calculated, each using a distinct cross-validation framework. For classification, LOOCV was performed across EEG data from all patients, and a threshold of ρ ≥ 0.5 was applied to assign class labels. For correlation analysis, LEAPD indices were recomputed specifically for the 22 deceased patients using a separate leave-one-out procedure. In each iteration, data from one deceased subject were excluded, and the hyperplanes were trained on data from the remaining 21 deceased and all 22 living patients. The resulting LEAPD indices ranged between 0 and 1 and were not thresholded. These LEAPD indices were then correlated with the number of days from EEG acquisition to date of death for each of the deceased participants. LEAPD indices with a higher value were interpreted as indicating shorter survival duration, enabling the evaluation of LEAPD as a continuous prognostic biomarker in PD.

### Performance Metrics

*Classification performance was evaluated using accuracy, sensitivity, specificity, and the area under the receiver operating characteristic curve (AUC). For correlation analyses, Spearman’s rank correlation coefficient p and corresponding p-values were computed. All metrics were calculated independently for each EEG channel or channel combination as relevant*.

### Data Availability

Deidentified data for each patient, along with essential analysis codes and detailed results, will be made available post-publication at: http://narayanan.lab.uiowa.edu and OpenNeuro.

## Results

### Patient Characteristics

Table 1 summarizes the demographic and clinical characteristics of the 94 PD patients that participated in this study, comparing the “living” (n = 72) and “deceased” (n= 22) groups. On average, the Deceased group was older (median [range]: 72 [64–82] vs. 68 [57–82] years, *p* = 0.004) and had more advanced disease severity, evidenced by higher Hoehn-Yahr stage (2.5 [1–6] vs. 1.8 [0–4], *p* = 0.002) and greater motor impairment, as assessed by UPDRS III (17.5 [4–28] vs. 12 [1–26], *p* = 0.0004). Cognitive function was also significantly lower in the deceased group (MoCA: 23 [16– 28] vs. 26 [11–30], *p* = 0.009). No significant differences were observed in disease duration in living group vs the deceased group (4 [0–24] vs. 5 [1–13] years, *p* = 0.15), levodopa equivalent daily dose (LEDD) (935.7 [300–2075] vs. 798.8 [150–1700] mg/day, *p* = 0.18), or sex distribution (72.7% vs. 65.3% male, *p* = 0.53).

**Table 1.** Clinical and demographic comparisons between Living (n = 72) and Deceased (n = 22) PD patients in this study. Continuous variables are reported as median [range]; *p* values are from Mann-Whitney U tests (for continuous/ordinal variables) or Chi-square tests (for sex distribution).

| **Measure** | **Living (N=72)** | **Deceased (N=22)** | ***p* value** | **Test Used** |
| --- | --- | --- | --- | --- |
| **Age (years)** | 68 [57–82] | 72 [64–82] | 0.004* | Mann-Whitney U |
| **Sex (% male)** | 65.3 | 72.7 | 0.53 | Chi-square |
| **Disease duration (years)** | 4 [0–24] | 5 [1–13] | 0.15 | Mann-Whitney U |
| **Hoehn-Yahr stage** | 2 [0–4] | 2.5 [1–6] | 0.002* | Mann-Whitney U |
| **UPDRS III (motor)** | 12 [1–26] | 17.5 [4–28] | 0.0004* | Mann-Whitney U |
| **LEDD (mg/day)** | 750 [150–1700] | 950 [300–2027] | 0.18 | Mann-Whitney U |
| **MoCA** | 26 [11–30] | 23 [16–28] | 0.009* | Mann-Whitney U |
| **Time to death (months)** | - | 45 [9–79] | - | - |

\* indicates $p < 0.05$ .

### Initial Classification and Truncation Analysis

Single-channel LEAPD indices were first evaluated using a balanced dataset of 44 PD patients (22 deceased, 22 living) using LOOCV. Classification performance was assessed independently for each EEG channel, with most channels achieving 100% accuracy and the lowest observed accuracy being 93.33%, indicating strong discriminative power (Figure 1e).

To evaluate classifier robustness under reduced data conditions, a truncation analysis was performed on the 44-subject dataset. EEG recordings were truncated to retain 90%, 66%, or 50% of their original length, and classification accuracy was obtained without re-optimizing LEAPD parameters. The top 5 performing channels (CP4, PO4, FCz, P2, and P5) maintained strong classification performance across all truncation levels, as shown in the accuracy heatmap (Figure 1f). For instance, FCz achieved 100.0%, 97.7%, 84.1%, and 81.8% accuracy at 100%, 90%, 66%, and 50% data retention, respectively, while P5 sustained performance with 100%, 100%, 81.8%, and 72.7% accuracy.

To further assess the impact of data truncation on individual channel discriminability, LEAPD indices for P5 were analyzed across truncation levels. As shown in Figure 2a, the distributions between the deceased and living groups remained distinct even as the EEG data were reduced. The corresponding ROC curves (Figure 2b) show AUCs of 1.00 (100% and 90% truncation), 0.89 (66%), and 0.74 (50%), demonstrating the robustness of the classifier under data constraints.

**Figure 2.**
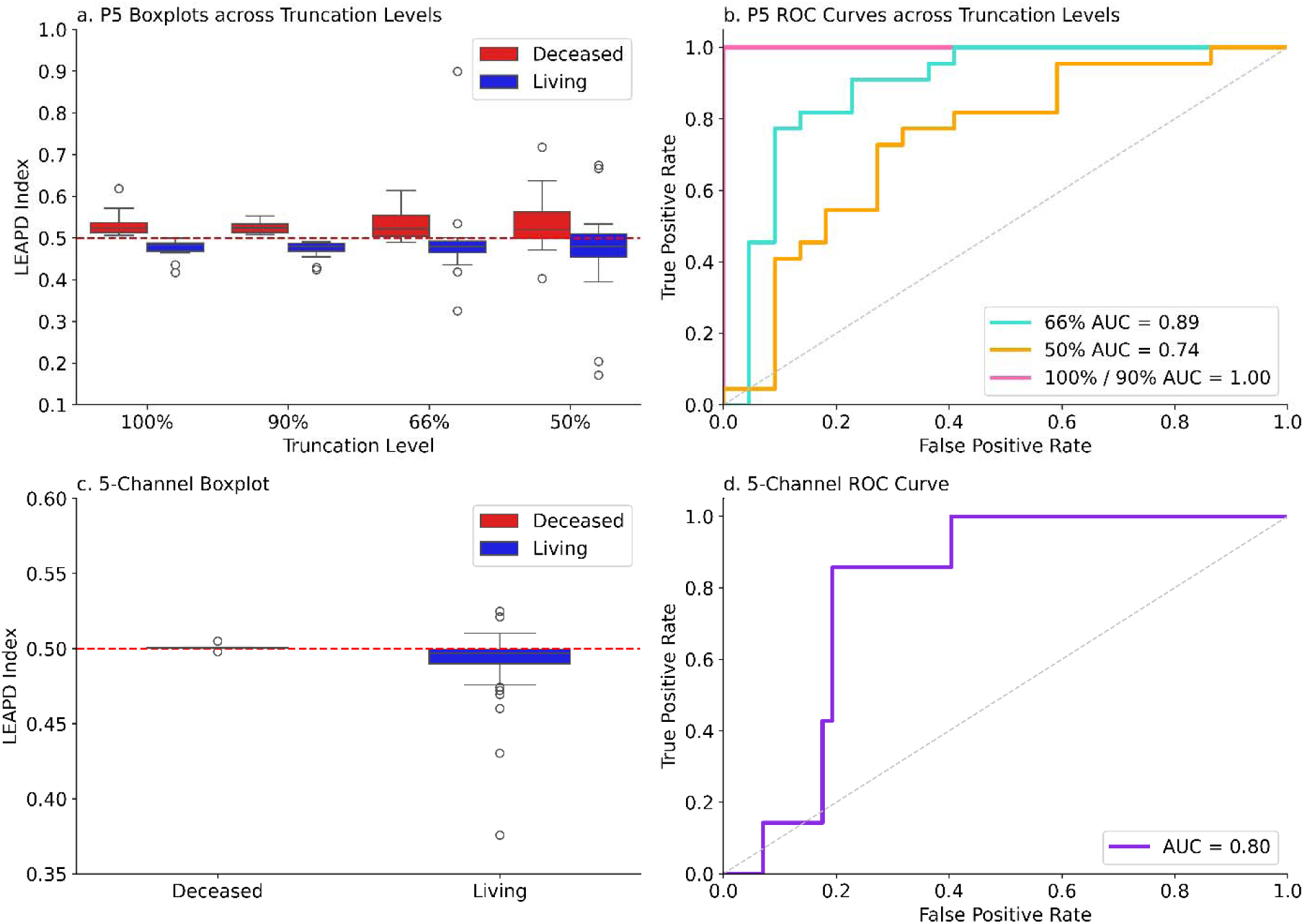
LEAPD classification performance under truncation and multi-channel setup. **(a)** Boxplots of LEAPD indices for deceased and living participants using channel P5 across varying data truncation levels, showing preserved group separation. **(b)** Receiver Operating Characteristic (ROC) curves for P5 classification at each truncation level, with performance measured by area under the ROC (AUC). **(c)** Boxplot of LEAPD index distributions for the 5-channel classifier, showing enhanced separation between groups. **(d)** ROC curve for the 5-channel classifier evaluated using out-of-sample data, achieving an AUC of 0.80.

### Out-of-Sample Classification: 7-vs-7 Testing

Separately, LEAPD parameters were optimized using data from a distinct training cohort of 30 PD patients (15 deceased, 15 living). These optimized parameters were then applied for out-of-sample validation on data from an independent cohort of 64 PD patients (7 deceased, 57 living). Balanced comparisons were achieved by generating 10,000 random subsets of data from 7 living participants to compare against the 7 deceased participants.

A 5-channel combination (CP2, FC5, F4, PO4, CP4) was evaluated across all 10,000 randomized 7-vs-7 comparisons, yielding a mean classification accuracy of 83% with a standard deviation of 7%, confirming the robustness and reproducibility of the LEAPD-based mortality classifier. As shown in Figure 2c, the LEAPD indices remained well separated between the deceased and living groups. The corresponding ROC curve in Figure 2d demonstrates strong discriminative performance, with an AUC of 0.80 for the 5-channel classifier.

### Correlation Between LEAPD Indices and Survival Duration

To assess whether the LEAPD index reflects proximity to death, we performed an LOOCV analysis restricted to data from the 22 deceased patients. For each subject, we constructed deceased and living hyperplanes using the remaining 21 deceased and 22 living patients, respectively, and calculated the LEAPD index for the held-out subject. This process was repeated independently for each EEG channel. Spearman’s rank correlation was computed between LEAPD indices and the number of days from recording EEG to death. Most EEG channels exhibited strong negative correlations, supporting the hypothesis that higher LEAPD indices are associated with shorter survival durations. The 5 channels with the strongest negative correlations were F4 (ρ = –0.862, *p* = 1.49 × 10^⁻^), AFz (ρ = –0.832, *p* = 1.24 × 10^⁻^), AF7 (ρ = –0.811, *p* = 4.93 × 10^⁻^), Cz (ρ = –0.798, *p* = 1.11 × 10⁻), and AF4 (ρ = –0.796, *p* = 1.26 × 10^⁻^).

Notably, even the weakest correlation across all channels (observed at TP7) was still substantial (ρ = –0.703), and its age-adjusted correlation further improved to ρ = –0.756. In the out-of-sample correlation analysis, channel C1 yielded ρ = –0.821, and P6 showed ρ = –0.642. These findings suggest that the LEAPD index may serve not only as a binary classifier but also as a continuous prognostic indicator of survival in PD. After adjusting each correlation value separately for age, UPDRS III score, and MoCA score, LEAPD correlations remained significant.

### Comparative Analysis

We compared our findings with previous studies on mortality prediction and biomarker development in PD. Table 2 summarizes existing methodologies, predictive markers, and performance metrics. Compared to traditional motor, cognitive, and biochemical predictors, LEAPD offers a noninvasive and objective EEG-based method with competitive accuracy, robustness to data reduction, and strong generalizability. These results further support the potential clinical utility of EEG-based mortality prediction using the LEAPD framework.

**Table 2.**
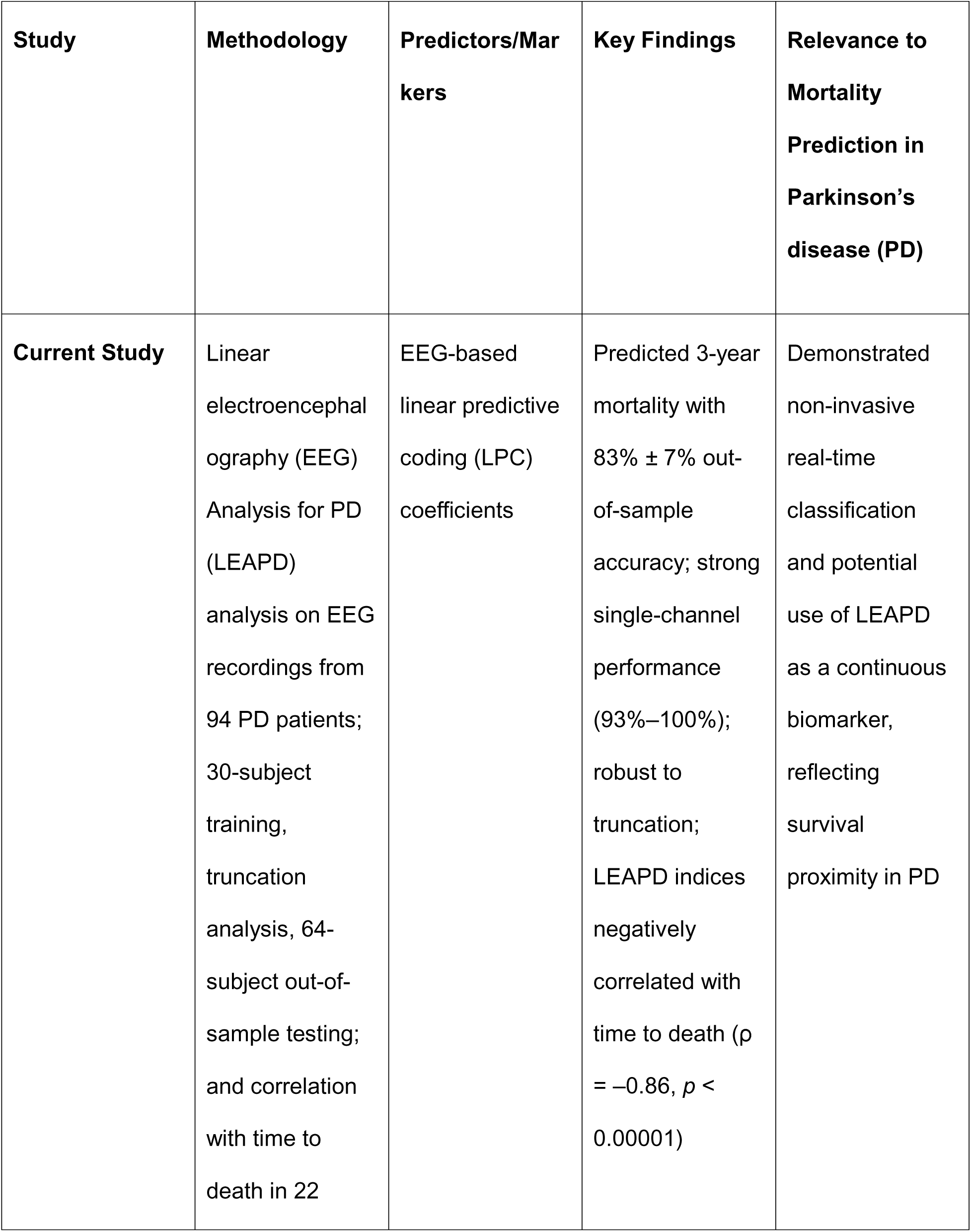
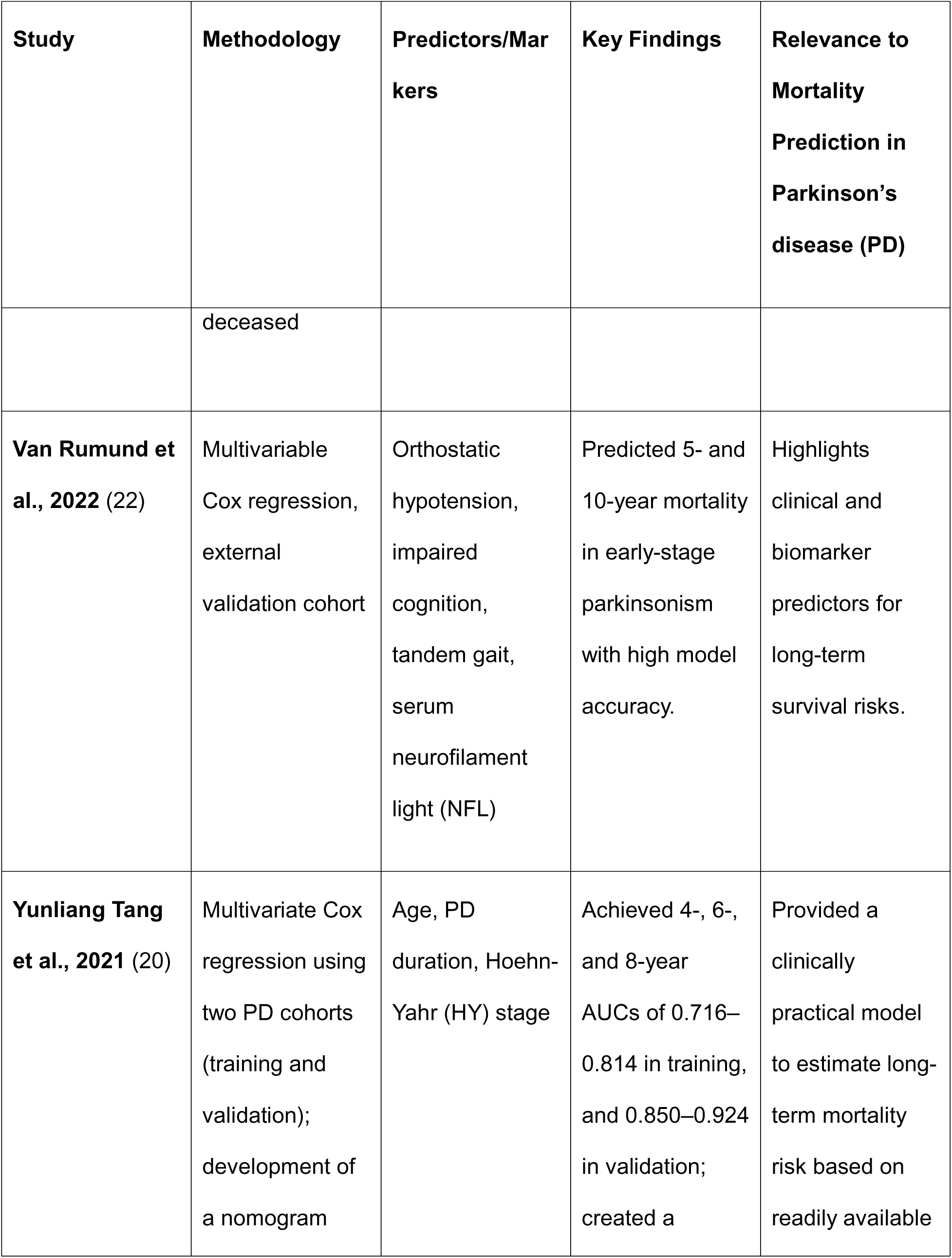
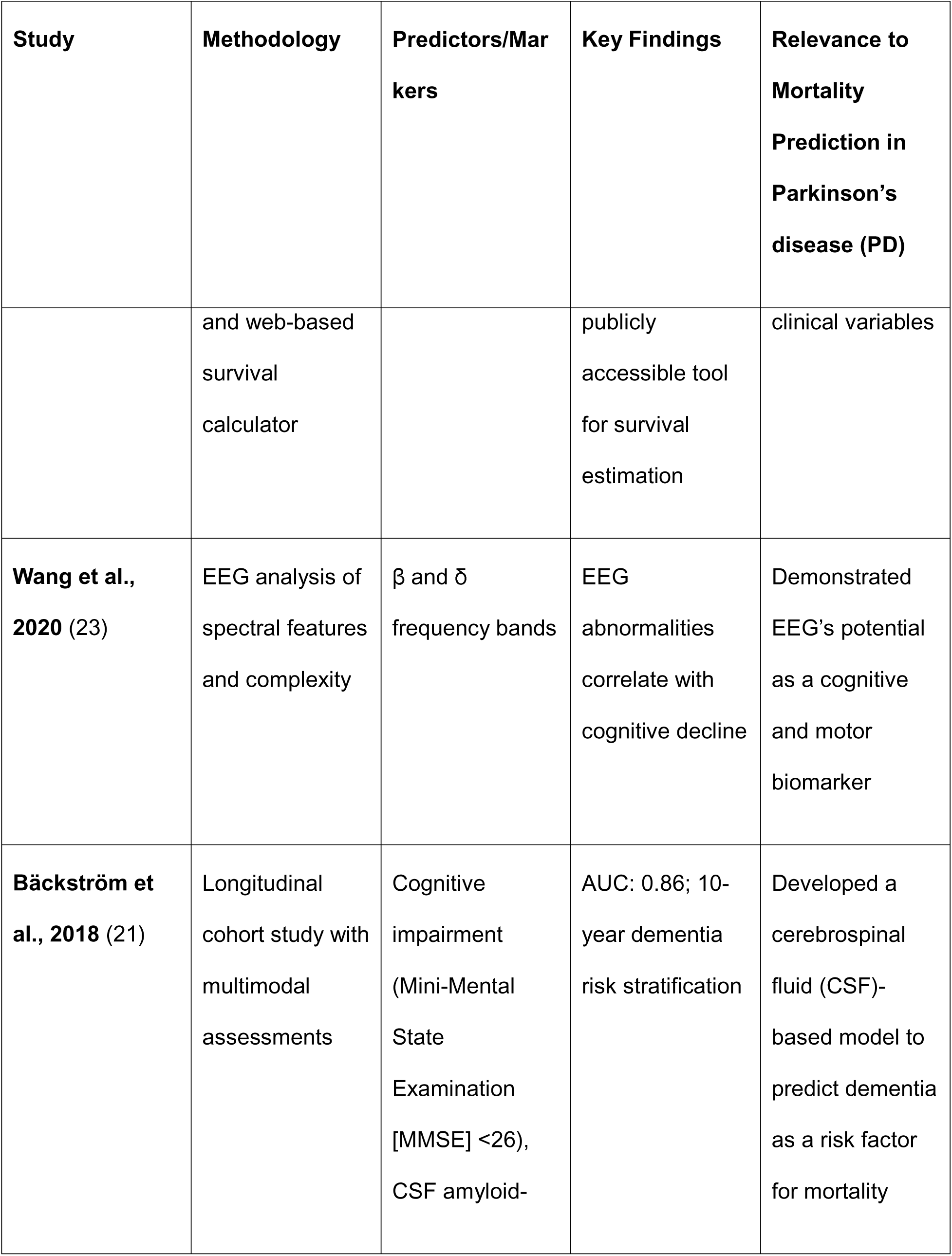
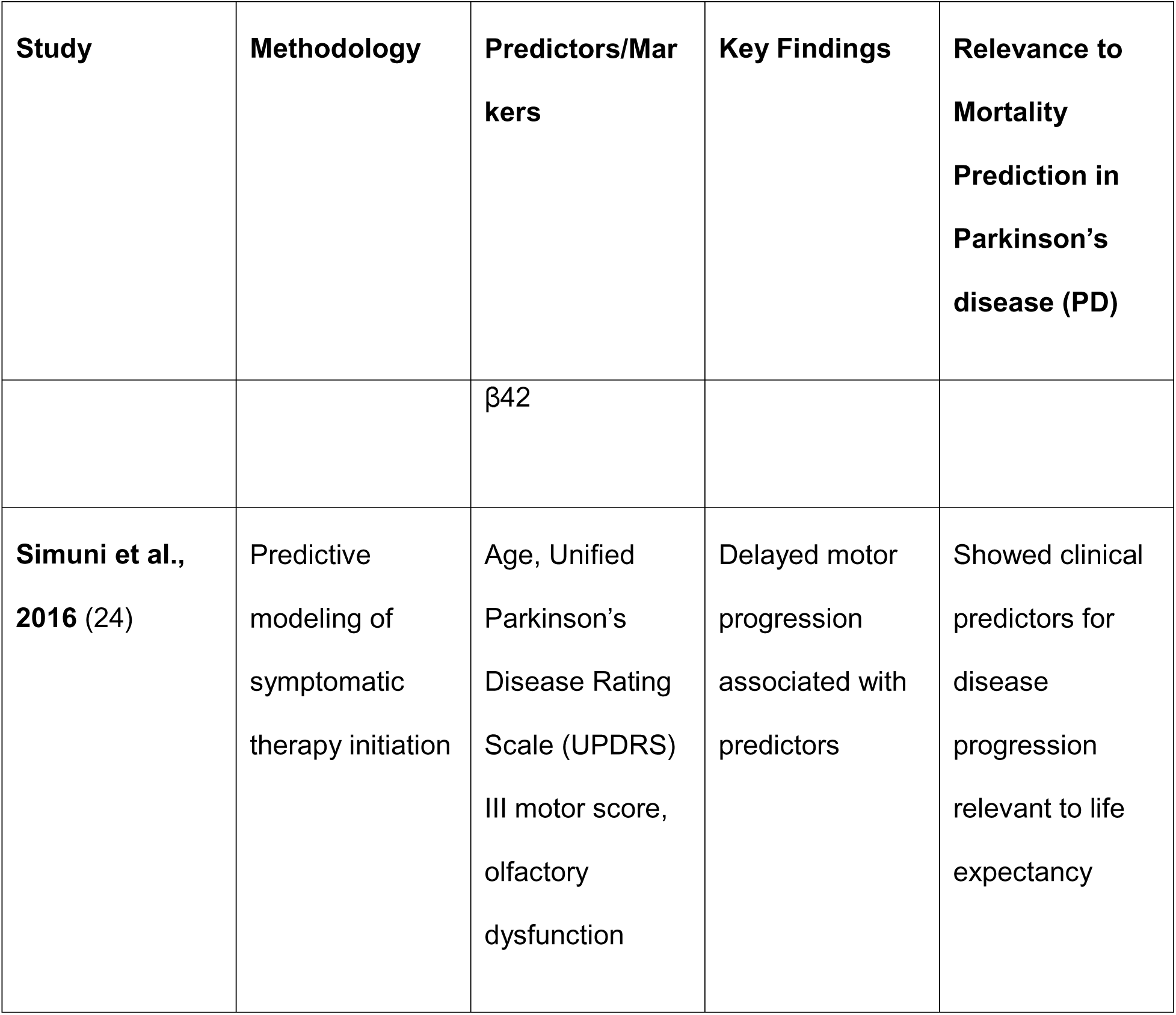
Comparison of this study with existing research on mortality prediction in Parkinson’s disease. This table compares methodologies, predictors, and outcomes from both this study and related research on mortality prediction in PD highlighting non-invasive EEG-based biomarkers vs traditional clinical or biochemical markers.

## Discussion

This study investigated the Linear Predictive Coding EEG Algorithm for Parkinson’s disease (LEAPD) as a potential non-invasive biomarker for mortality prediction in PD. Using a two-stage analysis, we first demonstrated strong single-channel classification performance in a balanced cohort of 44 PD patients (22 deceased, 22 living) using LOOCV and confirmed classifier robustness through truncation analysis under reduced data conditions. We then validated the generalizability of the LEAPD framework through an out-of-sample evaluation involving a separately optimized training cohort of 30 PD patients (15 deceased, 15 living) and an independent testing cohort of 64 PD patients (7 deceased, 57 living), using 10,000 randomized 7-vs-7 comparisons.

LEAPD achieved strong classification performance across both stages, with the optimal 5-channel combination (CP2, FC5, F4, PO4, CP4) yielding a mean out-of-sample accuracy of 83% ± 7%. In addition to its classification performance, LEAPD indices demonstrated a strong negative correlation with time to death among deceased PD patients. This finding supports the potential of the LEAPD index as a continuous prognostic biomarker that may reflect not only mortality risk but also the relative proximity to death. Even after adjusting for age, UPDRS III score, and MoCA score, the LEAPD index was still highly correlated with time to death, further emphasizing its utility as a mortality predictor. Such a relationship enables the LEAPD index to offer more nuanced clinical information beyond binary classification, potentially informing patient monitoring and care planning strategies.

These results highlight the promise of LEAPD as a computationally efficient, scalable, and clinically feasible tool for mortality risk assessment in PD. By encoding EEG time-series data through LPC vectors and leveraging simple single-or multi-channel classifiers, LEAPD provides an objective framework that could facilitate earlier identification of high-risk patients and support clinical decision-making. Beyond binary classification, the significant correlation between LEAPD indices and time to death suggests that the LEAPD index may also serve as a continuous marker of mortality risk. Its non-invasive nature and minimal channel requirements further position it as an attractive alternative to cerebrospinal fluid (CSF) or serum-based biomarkers, which are often limited by cost, invasiveness, and patient compliance.

Despite these strengths, two limitations must be acknowledged. The cohort sample size may limit the generalizability of the findings. Although this study represents one of the largest prospective EEG datasets focused on PD mortality classification to date, larger and more diverse populations will be needed to confirm the robustness of these results. Additionally, variability in EEG collection conditions—such as electrode placement, participant movement, and medication status—may affect reproducibility across different clinical settings. Standardization of EEG acquisition protocols will be critical for future validation efforts.

Overall, this work underscores the novelty and potential of EEG-based approaches for mortality prediction in PD—a focus that remains underexplored in prior research. While methods such as CSF biomarker analysis provide valuable insights, they often involve invasive procedures and are resource-intensive (20,21). LEAPD offers a non-invasive, computationally efficient, and clinically scalable alternative, capable of real-time classification. Moreover, its significant correlation with survival duration suggests that LEAPD may also serve as a continuous prognostic biomarker, enabling more nuanced assessment of mortality risk in PD.

## Data Availability

Deidentified data and analysis code will be made publicly available upon publication via the Narayanan Lab and OpenNeuro.

http://narayanan.lab.uiowa.edu

## Acknowledgments

This study was supported by the National Institutes of Health under grants R01NS100849-01A1 and 1RF1NS127809-01A1. The National Institute of Neurological Disorders and Stroke (NINDS) was not involved in the study’s design, data collection, analysis, interpretation, or manuscript preparation. We sincerely thank the staff of the Department of Neurology at the University of Iowa for their support and assistance throughout this research.

We thank Heather Widmayer of the Scientific Editing and Research Communication Core at the University of Iowa Carver College of Medicine for her editorial assistance. We thank Dr. Md Fahim Anjum for clarifying the implementation of the LEAPD algorithm.

## Authors’ Roles

SJ, NSN, SD, and EYU conceptualized the work. SJ and PM implemented the algorithm. AIE and JTH collected the data. SJ wrote the paper, and all authors edited the paper.

## Financial Disclosures of All Authors

None.

## Notes

### Competing Interest Statement

The authors have declared no competing interest.

### Author Declarations

The Institutional Review Board of the University of Iowa gave ethical approval for this work (IRB# 201707828).

