## Supplementary Materials: Tables and Methods for "Linear predictive coding electroencephalography algorithms predict Parkinson’s disease mortality using out-of-sample tests": Hyperparametersfor outofsampleaccuracy.docx

**Table A2.** Chosen Channels and Their Top-Performing Parameters

| Channel | Parameters | | |
| --- | --- | --- | --- |
|  | Filter range (Hz) | LPC order | Number of components |
| CP2 | 0.1 – 44 | 9 | 4 |
| FC5 | 0.1 – 6 | 3 | 2 |
| F4 | 0.3 – 40 | 9 | 1 |
| PO4 | 0.1 – 17 | 3 | 1 |
| CP4 | 0.1 – 79 | 10 | 1 |

Optimal Parameter Values for Selected Channels in Single and 5-Channel Classification
