## Supplementary Materials: Tables and Methods for "Linear predictive coding electroencephalography algorithms predict Parkinson’s disease mortality using out-of-sample tests": LEAPD Math.docx

**Mathematical Explanation of the LEAPD Approach**

1. **Feature Extraction Using LPC**

Linear Predictive Coding (LPC) algorithm encodes the EEG time series into coefficients that describe the signal's power spectral density (PSD) [1]. Using Burg's method [1], [2], [3], [4], the LPC coefficients are computed by minimizing the energy of both forward and backward prediction errors recursively.

Let the EEG time series consist of
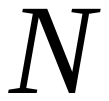
samples, denoted as
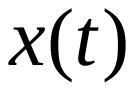
 for
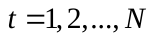
. The
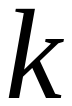
-th order LPC coefficients,
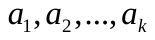
, are calculated by minimizing the prediction error:

- Forward Prediction Error:

| 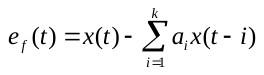 | (1) |
| --- | --- |

- Backward Prediction Error:

| 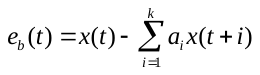 | (2) |
| --- | --- |

- Total Prediction Error:

| 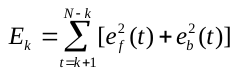 | (3) |
| --- | --- |

Burg's method solves this optimization problem iteratively for
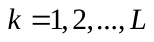
, where
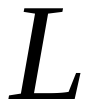
 is the chosen LPC order. This process generates
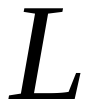
LPC coefficients that are used as feature vectors for classification.

2. **Principal Component Analysis for Classification**

The LPC feature vectors of Parkinson's disease (PD) and control subjects form distinct subspaces. Principal Component Analysis (PCA) is used to identify these subspaces by capturing the variance in the feature vectors [1], [5].

- **Data Matrix Construction:** Let
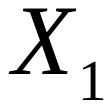
 and
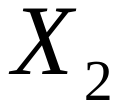
 represent the feature matrices of
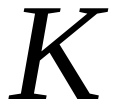
deceased PD subjects and
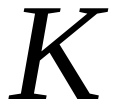
living PD subjects, respectively:

| 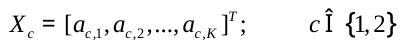 | (4) |
| --- | --- |

- **Bias Vector Calculation:**

| 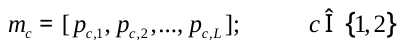 | (5) |
| --- | --- |

Where each
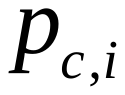
is the mean of the
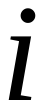
-th column of
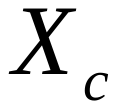
.

3. **Single-Channel Classification**

To process single-channel EEG data, several steps are undertaken:

- **Energy Normalization:** The EEG signal is normalized to unit energy:

| 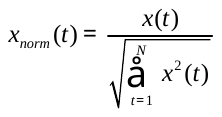 | (6) |
| --- | --- |

- **Bandpass Filtering:** A 6th-order Butterworth bandpass filter is applied to retain relevant frequency components [1].
- **Feature Extraction:** LPC coefficients are derived from the filtered EEG signal and used as feature vectors for classification [1].

4. **Subspace Formation and Classification**

Principal Component Analysis (PCA) creates distinct subspaces for PD and control subjects based on their LPC feature vectors.

- **Data Organization**: Feature matrices for deceased and living subjects are constructed:
- **Mean Removal**: Bias vectors
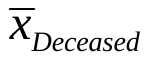
 and
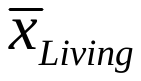
 are subtracted to center the data:

| 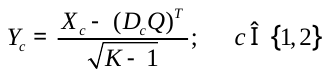 | (7) |
| --- | --- |

- Where
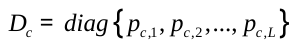

-
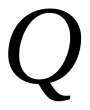
 is an
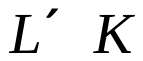
matrix with all elements equal to 1
- **PCA via SVD**: Singular Value Decomposition (SVD) is applied:

| 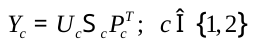 | (8) |
| --- | --- |

- Where
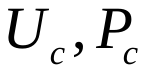
are orthogonal matrices
-
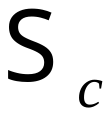
are diagonal matrices containing singular values in decreasing order

5. **Classification Using the LEAPD Index**

A new subject's LPC feature vector,
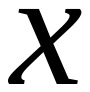
, is projected onto the deceased and living hyperplanes. The distances from these projections are used to compute the LEAPD index.

- **Projection and Distance Calculation:**

| 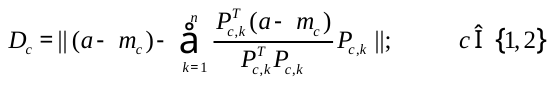 | (9) |
| --- | --- |

- **LEAPD Index:** The LEAPD index,
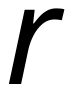
, is calculated as:

| 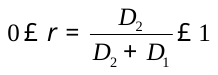 | (10) |
| --- | --- |

- If
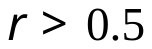
, the subject is classified as deceased.
- If
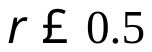
, the subject is classified as livings.

6. **Parameter Optimization**

The classifier parameters, such as the LPC order
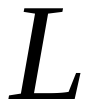
 and the number of principal components
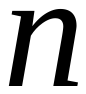
, are optimized through exhaustive search to maximize classification accuracy. Bandpass filters in the 0.5-10 Hz range are applied to the EEG signals to capture the most relevant PSD features [1], [6]. The LPC order is varied (e.g.,
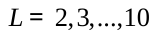
) to identify the best-performing configuration [1], [7].

7. **Validation and Real-Time Application**

Out-of-sample test is used to evaluate performance. The LEAPD algorithm's computational simplicity ensures low latency, making it suitable for real-time applications [1].
