## Supplementary Materials: Tables and Methods for "Linear predictive coding electroencephalography algorithms predict Parkinson’s disease mortality using out-of-sample tests": Table for single channel LOOCV accuracy.docx

**Table A1.** Single-Channel LOOCV Accuracy Results

| Channel’s name | LOOCV Accuracy (%) |
| --- | --- |
| C3 | 93.182 |
| F7 | 95.455 |
| CP1 | 95.455 |
| PO4 | 95.455 |
| P6 | 95.455 |
| P3 | 97.727 |
| P7 | 97.727 |
| O2 | 97.727 |
| P4 | 97.727 |
| P8 | 97.727 |
| CP6 | 97.727 |
| Cz | 97.727 |
| F4 | 97.727 |
| F1 | 97.727 |
| FT7 | 97.727 |
| C5 | 97.727 |
| CP3 | 97.727 |
| PO7 | 97.727 |
| PO3 | 97.727 |
| C2 | 97.727 |
| Fp1 | 100 |
| Fz | 100 |
| F3 | 100 |
| FC5 | 100 |
| FC1 | 100 |
| T7 | 100 |
| CP5 | 100 |
| O1 | 100 |
| Oz | 100 |
| CP2 | 100 |
| C4 | 100 |
| T8 | 100 |
| FC6 | 100 |
| FC2 | 100 |
| F8 | 100 |
| Fp2 | 100 |
| AF7 | 100 |
| AF3 | 100 |
| AFz | 100 |
| F5 | 100 |
| FC3 | 100 |
| C1 | 100 |
| TP7 | 100 |
| P1 | 100 |
| P5 | 100 |
| POz | 100 |
| PO8 | 100 |
| P2 | 100 |
| CPz | 100 |
| CP4 | 100 |
| TP8 | 100 |
| C6 | 100 |
| FC4 | 100 |
| FT8 | 100 |
| F6 | 100 |
| AF8 | 100 |
| AF4 | 100 |
| F2 | 100 |
| FCz | 100 |
